# Primary Care Providers Journey with OSA Care, Challenges and Strategies: A Qualitative Study

**DOI:** 10.64898/2026.05.15.26353339

**Authors:** Wanjae Cho, Michelle Cheng, Kenneth G. Blades, Oliver David, Willis H. Tsai, Marcus Povitz, Kerry A. McBrien, Maoliosa Donald, Sachin R. Pendharkar

## Abstract

**Purpose:** Obstructive sleep apnea (OSA) is a treatable chronic condition associated with significant health and societal consequences. Primary care providers (PCPs) often manage OSA with support from sleep specialists but face challenges navigating a complex system of care. By developing a Journey Map, we sought to identify factors influencing primary care OSA management and the associated PCPs’ perspectives and emotions.

**Methods:** Twenty-one Calgary-based PCPs were interviewed as part of a study evaluating a primary care management pathway for OSA. We used conventional content analysis, utilizing inductive coding to define journey phases and deductive coding via the Theoretical Domains Framework (TDF) to identify barriers and enablers. These were then mapped onto journey phases for OSA management to create a Journey Map.

**Results:** The Journey Map included five phases of OSA care. PCPs described feeling neutral during the Learning phase and expressed neutral to positive emotions during the Patient Encounter and Diagnosing OSA phases. In contrast, the Initial Treatment and Ongoing Management phases were associated with neutral to negative emotional experiences. Barriers included limited OSA-related training and education, unclear roles among provider groups, and low patient engagement. Enablers included accessible knowledge resources, a shared key role in OSA screening, and availability of sleep testing. Opportunities to enhance primary care OSA management were identified at each step.

**Conclusion:** This study identified several behavioural factors influencing PCP decision-making across the OSA care continuum. The Journey Map illustrates how high diagnostic confidence of PCPs shifts to escalating challenges and negative sentiment during treatment and long-term management of OSA.

## INTRODUCTION

Obstructive sleep apnea (OSA) is a prevalent and treatable chronic condition that is associated with significant health and societal impacts. A 2024 study estimated that approximately 28% of Canadian adults aged 45 to 85 have moderate to severe OSA [1]. Untreated OSA may increase the risk of hypertension, cardiovascular disease, dementia, and depression [2–4]. It is also associated with motor vehicle accidents, reduced productivity in the workplace, and increased healthcare and societal costs [5–9].

Although OSA treatment is cost-effective and leads to improved clinical outcomes [10–14], timely access to OSA management remains a challenge due to the increasing demand for sleep medicine services that outstrips sleep specialist capacity [15–17]. Given that prompt diagnosis and early initiation of OSA treatment are associated with improved therapy adherence and better patient outcomes [18], OSA management by primary care providers (PCPs) has been proposed [19–21]. Several studies have shown that OSA management in primary care settings yields comparable outcomes to specialist-led care while helping to reduce healthcare costs [22–26]. Moreover, patients often prefer to receive OSA care from their PCP [27], which aligns with broader evidence suggesting patients favour PCPs for managing chronic conditions [28, 29].

Primary care providers routinely manage chronic conditions like OSA [30, 31], but little is known about how they navigate OSA care delivery as "service users" within the current care model. In our prior qualitative analysis exploring patient and PCP perspectives, challenges related to care navigation and role clarity were identified as key gaps [27]. PCPs act as both the first point of contact for patients and central coordinators within a complex care system (Figure 1), which is a commonly adopted model of OSA care across Canada and globally [32–35]. As such, a deeper understanding of PCPs’ behaviours, experiences, and role perceptions is essential to strengthening their engagement and enhancing the effectiveness of PCP-led OSA care.

**Figure 1.**
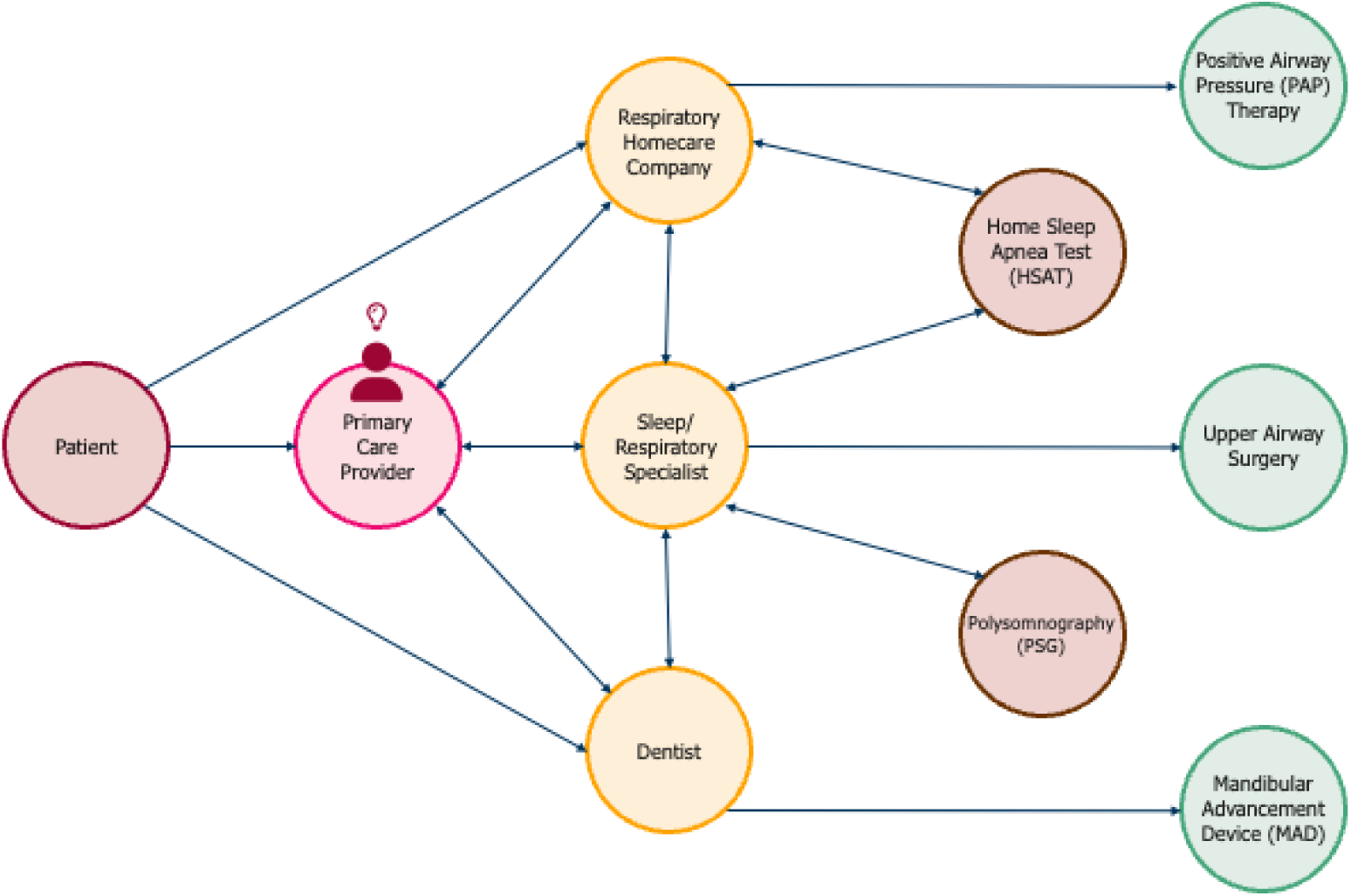
OSA Care Model in Alberta. The arrows illustrate the interplay between PCPs, patients, and other key partners involved in the diagnosis, management, and treatment of OSA in Alberta.

Journey mapping is widely used in behavioural sciences to provide a structured, visual representation of a service process from the perspective of the service user [36]. Applied to healthcare, it offers a detailed view of patient and provider experiences, interactions, and perspectives at each stage of disease management to identify gaps and target areas for intervention [37, 38]. This approach complements the Theoretical Domains Framework (TDF) [39], which integrates multiple behaviour change theories to identify the determinants of healthcare provider behaviour (Appendix 1).

This study had two primary objectives. First, we aimed to map the journey of PCPs in delivering care for OSA by capturing their experiences across the care continuum. Second, we sought to identify behavioural barriers and enablers at each phase of the journey using the TDF. By doing so, we established a foundation for designing targeted interventions to support sustained PCP engagement and improve the effectiveness of PCP-led OSA care.

## METHODS

### Study Design

We conducted a qualitative analysis of interview data from a study evaluating a clinical pathway to support primary care OSA management in Calgary, Canada. This study was approved by the University of Calgary Conjoint Health Research Ethics Board (Ethics ID: REB21-1978).

### Study Setting

In Alberta, patients with suspected OSA may be referred by their PCP to a sleep or respiratory specialist or for home sleep apnea testing (HSAT), which is performed by private respiratory homecare companies or either public or private sleep laboratories (Figure 1). Following diagnosis, PCPs may continue to direct treatment or refer the patient to a specialist, who may order in-lab polysomnography if indicated. Continuous positive airway pressure (CPAP) therapy is typically initiated through respiratory homecare companies staffed by respiratory therapists and polysomnographic technologists, while other therapies like mandibular advancement devices or upper airway surgery may be accessed through dentists or head and neck surgeons. Treatment is mostly privately funded with the exception of government funding programs for patients with low income or complex sleep-disordered breathing requiring non-invasive ventilation or supplemental oxygen.

### Study Participants

Primary care providers (family physicians and nurse practitioners) practicing in Calgary and surrounding area were recruited using advertisements in clinical and academic email newsletters distributed by primary care organizations, social media advertisements that were shared within provider networks, and through snowball sampling via study participants. Interviewees provided informed, written consent prior to participation and were provided a gift card to acknowledge their time and contributions.

### Data Collection

Semi-structured interviews were conducted between May and August 2024 by a coinvestigator with qualitative expertise and knowledge about OSA and the primary care pathway (KGB). Interviews followed an interview guide (Appendix 2) that was designed to gather perspectives on the pathway and explore a broad range of topics related to OSA care. The interview focused on: self-perceived role in managing OSA, perceptions of the primary care OSA clinical pathway [40], interactions with other individuals involved in OSA care, and enablers and barriers to OSA care in their clinical practice. We provided the pathway to participants to review in advance and as needed during interviews. Interviews were audio-recorded and transcribed verbatim using an AI-based transcription software, Rev (Austin, TX, USA) with subsequent review of recordings and transcripts by another investigator (MC) to verify accuracy. Data was collected and stored securely on cloud-based servers at the University of Calgary.

### Data Analysis

Two investigators with clinical (WC) and qualitative research (MC) experience independently familiarized themselves with the data. We acknowledged that the investigators’ differing viewpoints could influence data generation and analysis. Therefore, coding was done independently and subsequently through regular meetings and discussion consensus was achieved for all steps of the analytical process. All transcript data were stored and coded using NVivo software (version 12; QSR International, Burlington, MA, USA).

We employed a three-step analytic process to comprehensively examine PCPs’ experiences with OSA management. First, interview transcripts were inductively analyzed using conventional content analysis to derive the phases of the OSA care journey for PCPs [41]. Through iterative transcript review, excerpts reflecting key experiences and processes were coded and organized into preliminary categories, from which five journey phases were identified. Representative quotes were then mapped to each phase to construct the Journey Map. The resulting Journey Map visualized the care process from the PCP perspective, capturing both practical dimensions and emotional landscapes within OSA care delivery.[42, 43]

Second, we used a deductive approach to identify behavioural barriers and enablers at each journey phase using the 14 TDF domains. We identified key domains from the TDF at each journey phase using a summative content analysis approach [41]. Within each domain, coded data were categorized as a Barrier, Enabler, or labeled as ‘Both’ if encompassing elements of both categories across the journey phases. Domains were considered key if they met all the following criteria: the associated barrier or enabler was mentioned by more than two PCPs, there was a high frequency of specific beliefs within the domain, and there was conflicting or strongly held beliefs that could influence patient care and clinical outcomes.

Finally, thematic analysis following Braun and Clarke’s six-step method [44] was conducted to identify themes along the five phases of the Journey Map. All relevant TDF domains were organized into themes to highlight PCPs’ behaviours, patterns, and emotional responses related to OSA management.

## RESULTS

We analyzed interview transcripts from 21 PCPs (20 family physicians, one nurse practitioner). Eighteen (86%) participants identified as women and median (IQR) duration of practice was 7 (9) years. While all participants reviewed the primary care pathway prior to interviews, only 3 (14%) had previously used it. Irrespective of prior pathway use, participants discussed practice- and system-level factors influencing OSA care.

### OSA Care Journey Map

We identified five key phases in the Journey Map: Learning, Patient Encounter, Diagnosing OSA, Initial Treatment for OSA, and Ongoing OSA Management. Figure 2 illustrates the Journey Map and associated PCP experiences and behaviours.

**Figure 2.**
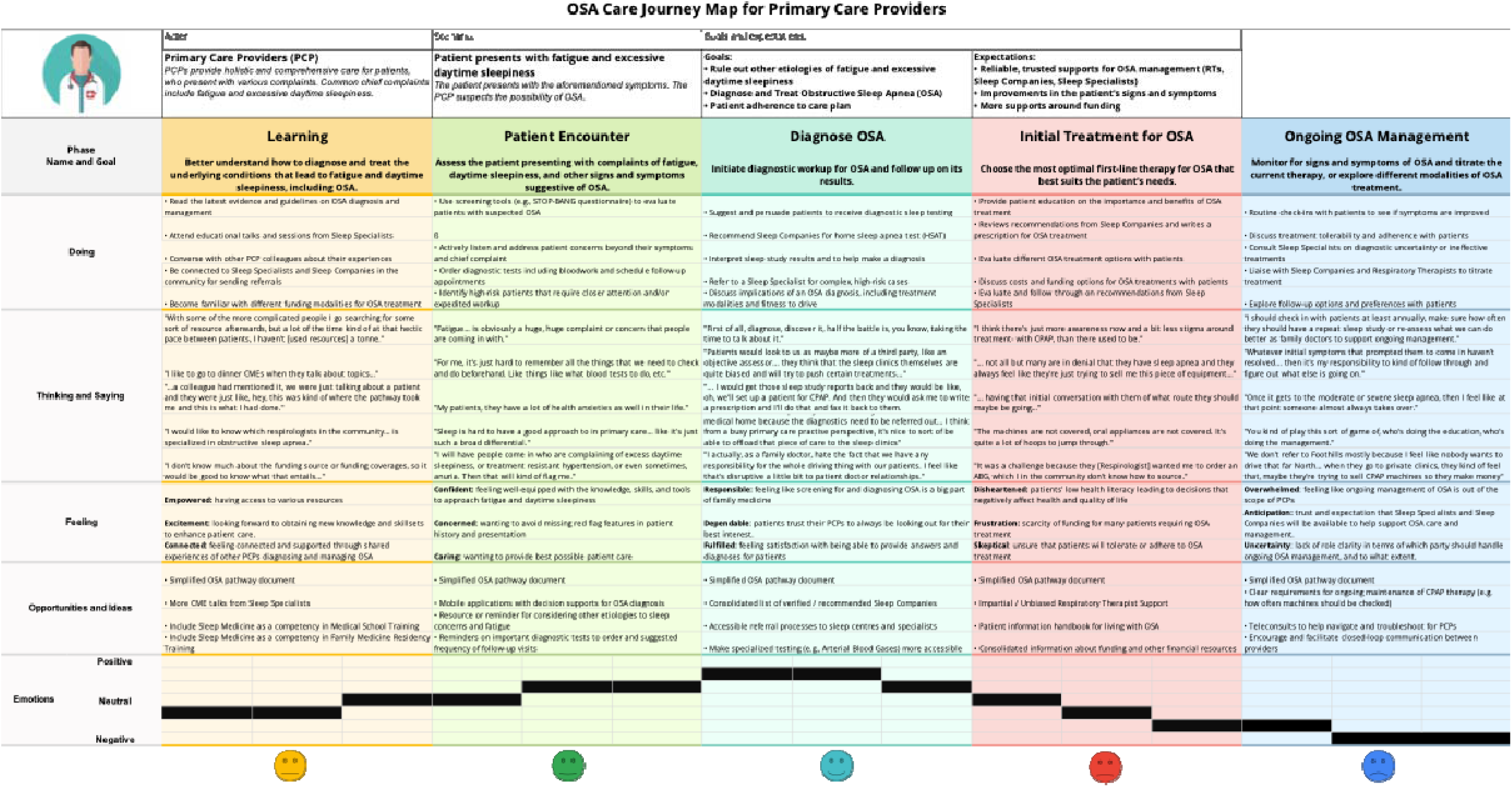
PCP OSA Care Journey Map. The OSA care journey for PCPs are depicted across five key journey phases:. ‘Doing’ – specific PCP actions and behaviours; ‘Thinking and Saying’ –thoughts, reflections, and verbalized experiences of PCPs;. ‘Feeling’ – emotional responses PCPs commonly experience; ‘Opportunities and Ideas’ –potential improvements, strategies, and creative suggestions to enhance OSA care and address existing barriers; ‘Emotions’ – dominant emotional tone ranging from negative ( ‘®’), neutral ( ‘@’), or positive ( ‘☺’).

The Journey Map revealed different roles, emotions, and tensions as PCPs navigated OSA care. Early in the process, during learning and patient encounters, PCPs expressed curiosity and motivation to expand their knowledge, though limited time and resources constrained engagement. Emotions were generally neutral to positive, supported by strong therapeutic relationships with patients. Confidence and professional identity peaked during the diagnosis phase, where PCPs described fulfillment in guiding patients through testing and acting as objective assessors. In contrast, treatment initiation and ongoing management were marked by greater uncertainty, frustration, and role ambiguity, which eroded confidence and fostered predominantly negative emotions. Overall, PCPs began their OSA care journey with interest and confidence. However, as care advanced, their engagement diminished due to the cumulative effects of a fragmented system of OSA care and competing demands.

Across all steps of the Journey Map, PCPs articulated a range of ‘Opportunities and Ideas’ that underscored the need for greater integration, clearer role delineation, and improved resourcing in OSA care. Proposed strategies included implementing standardized care pathways and strengthening communication channels with relevant partners. They also included providing clearer information on available diagnostic and treatment options, organized according to public and private funding pathways. Many participants emphasized the importance of shared-care models to clarify responsibilities between PCPs and other providers, as illustrated by one respondent: “I find we’re in our silo and the whole dental area is in their silo and there’s not much, there’s no communication really between dentists and doctors.” [PCP 3]. Others recommended embedding OSA-specific prompts or clinical decision-support tools within electronic medical records to mitigate cognitive load during time-constrained encounters.

### Key TDF Domains Across Journey Phases

Across the journey phases, several TDF constructs consistently shaped PCP engagement in OSA care (Table 1). Knowledge gaps and limited Environmental Context and Resources such as time, availability of diagnostic tests, and CPAP funding coverage, often constrained practice at all phases of the OSA care journey. Social Influences, such as collaborative relationships with other PCPs and specialists, motivated learning and enhanced confidence. Professional Role & Identity was most pronounced in diagnosis, where PCPs viewed themselves as key gatekeepers and central coordinators of OSA care. This sense of identity diminished in treatment and ongoing management, where uncertainty about scope and responsibilities was more evident. Beliefs about Capabilities varied, with some PCPs expressing confidence in initiating therapy while others deferred to specialists. Financial barriers and fragmented care pathways contributed to pessimism about long-term patient adherence, which in turn undermined PCPs’ perceptions of patients’ motivation to maintain follow-up. At the same time, Positive Reinforcement from observing patient improvements encouraged continued engagement. Taken together, PCPs expressed strong Professional Identity and Optimism early in the care journey, but their sustained engagement became tempered as barriers compounded across the continuum of care.

**Table 1.** Key TDF Domains Mapped to Each Journey Phase in PCP-led OSA Care.

| Journey Phase | Key TDF Domain | Significance |
| --- | --- | --- |
| Learning | Knowledge | PCPs highlighted both knowledge gaps and the value of education in recognizing OSA-related presentations. |
|  | Social Influences | Peer recommendations, professional norms, and interactions with sleep specialists or respiratory homecare companies influenced PCPs' motivation to learn more about OSA as a relevant educational and clinical priority. |
|  | Goals (I want to) | PCPs expressed internal motivation to improve patient outcomes in OSA care, which led to deeper engagement in OSA-related knowledge acquisition through various resources and specialists. |
| Patient Encounter | Memory, Attention, and Decision Processes | PCPs relied on clinical pathways and decision aids but also reported difficulty remembering all necessary steps during patient assessments. |
|  | Environmental Context and Resources | Since fatigue is a frequently reported chief complaint in primary care, it naturally aligned with opportunities for OSA screening. However, the heavy clinical workload often limited PCPs' ability to conduct a comprehensive assessment and workup. |
|  | Beliefs About Capabilities | Many PCPs reported feeling well-equipped and confident in managing patients presenting with fatigue and excessive daytime sleepiness. |
| Diagnosing OSA | Environmental Context and Resources | PCPs highlighted that timely access to diagnostic tests and resource constraints, including patient financial barriers, significantly influenced the diagnostic process. |
|  | Social/Professional Role and Identity | Many PCPs shared a self-perception as key players in screening for diseases and chronic conditions, including OSA. |
|  | Skills | PCPs are skilled in screening for concerning symptoms and initiating appropriate diagnostic workup and referrals. |
| Initial Treatment for OSA | Beliefs About Capabilities | Providers varied in their confidence regarding OSA treatment decisions. Some felt comfortable initiating therapy, while others felt this was outside their scope. |
|  | Environmental Context and Resources | The scarcity of funding for OSA treatment made it challenging for PCPs to initiate initial treatment for many patients. |
|  | Knowledge | Some primary care providers (PCPs) reported difficulty accessing information on where and how to initiate adjunctive treatments for OSA, such as oxygen therapy. |
| Ongoing OSA Management | Social/Professional Role and Identity | PCPs frequently reflected on whether long-term OSA management was within their scope of practice. Some saw themselves as key ongoing care providers, while others viewed OSA management as the responsibility of sleep specialists. This variability in role perception influenced their engagement in follow-up and care coordination. |
|  | Optimism | PCPs shared their perception that there appears to be more awareness about OSA in the general population and less stigma towards treatment. However, many expressed pessimistic views towards patient adherence to treatment due to prohibitive costs. |
|  | Emotion | When treatments for OSA are effective for patients and they see improvements in symptoms, PCPs were positively reinforced to support ongoing OSA treatment. Conversely, challenges such as unclear role responsibilities, high treatment costs, and prolonged wait times for referrals and diagnostic procedures served as negative |
|  |  | reinforcements, discouraging sustained engagement in OSA care. |

### Themes from Journey Phases

Bringing together the Journey Map and TDF analyses, three overarching themes emerged across OSA care journey phases: influence of systemic barriers, provider roles and responsibilities, and professional relationships and confidence (Table 2). Systemic barriers consistently constrained PCPs’ confidence and ability to provide comprehensive care across all phases of OSA care delivery. Participants described deficiencies in formal education on sleep medicine during their medical training, leaving many feeling underprepared to evaluate and manage OSA. These knowledge gaps were compounded by the environmental context of primary care, where short appointment times, limited diagnostic access, and inadequate funding for CPAP constrained practice and left little opportunity for PCPs to seek resources or engage in extended discussions about sleep-related symptoms. These barriers influenced PCPs’ beliefs about capabilities, with many expressing confidence in identifying OSA but feeling less assured when initiating treatment or managing long-term follow-up.

**Table 2.** Journey Phases and Associated Themes.

| **Themes** | **Subthemes** | **Supporting Quotes** | **Applicable Journey Phases** |
| --- | --- | --- | --- |
| **Influence of Systemic Barriers** | Gaps in formal education on OSA during medical training | "I would like to see a little bit of more education around like sleep disorders or like even just like insomnia. Just because I feel like again, we don't get a lot of education around it..." [PCP 6] | Learning, Initial Treatment for OSA, Ongoing OSA Management |
|  |  | "I have no idea what to do with a [CPAP] machine. I know it's got a bunch of buttons and that's about it." [PCP 2] |  |
|  | Time constraints of a busy primary care practice | "With some of the more complicated people I go searching for some sort of resource afterwards, but a lot of the time kind of at that hectic pace between patients, I haven't [used resources] a tonne." [PCP 1] | Learning, Patient Encounter, Ongoing OSA Management |
|  |  | "I think it's the different levels of tests that is most confusing... I, as a family doctor, would have to go and do some Google searching as to who offers what and what HSAT is exactly." [PCP 17] |  |
|  | Scarcity of funding sources and coverage | "I don't know much about the funding source or funding coverages, so it would be good to know what that entails..." [PCP 8] | Initial Treatment for OSA, Ongoing OSA Management |
|  |  | "I find a lot of my patients don't follow through on the CPAP. Funding is a big barrier for them, and I think just, for a lot of patients, even just convincing them that they need CPAP is also a barrier for them." [PCP 18] |  |
| **Provider Roles and Responsibilities** | Unclear scope of OSA care between partners | "I think what's tough about this care is like you can send somebody for a test and not know who they're gonna see and who they're going to. Like is it a physician? Is it someone who's gonna manage things? Are they gonna send things back to me to manage, will they look at the other symptoms?" [PCP 15] | Initial Treatment for OSA, Ongoing OSA Management |
|  |  | "I have no idea how to work your CPAP machine. Is that something that the PCP should be doing?" [PCP 14] |  |
|  | Ongoing patient management as PCPs' responsibility | "Whatever initial symptoms that prompted them to come in haven't resolved... then it's my responsibility to kind of follow through and figure out what else is going on." [PCP 12] | Ongoing OSA Management |
|  |  | "I mean certainly like the lifestyle modification piece sort of falls under general practice of family medicine." [PCP 17] |  |
|  | Therapeutic relationship and rapport between PCPs and patients | "Sometimes if it's a care pathway I'm particularly familiar with, and I know the patient's really anxious or they're concerned, I sometimes work it through with the patient... and some patients really like that." [PCP 13]<br>"[Telling patients that] you need a CPAP is a very easy conversation for me to have." [PCP 20] | Patient Encounter, Diagnosing OSA, Initial Treatment for OSA |
| Professional Relationships and Confidence | Shared self-perception of PCPs as key players in OSA screening | "First of all, diagnose, discover it, half the battle is, you know, taking the time to talk about it." [PCP 11]<br>"I have more of a role in the diagnostic side... just having that initial conversation with them of what route they should maybe be going." [PCP 1] | Patient Encounter, Diagnosing OSA |
|  | Adjunct treatments for OSA perceived as beyond PCPs' scope of practice | "Like when [respiratory homecare companies] send a thing back saying you could try the oral appliance versus the CPAP... I've never used the oral appliance." [PCP 18]<br>"For orthodontics and dental stuff, I just tell them talk to your dentist or orthodontist. Like you know, jaw issues and bite issues... they're comfortable dealing with sleep apnea." [PCP 4] | Initial Treatment for OSA, Ongoing OSA Management |
|  | Mutualistic relationships between PCPs and sleep specialists/respiratory homecare companies | "... I would get those sleep study reports back and [respiratory homecare companies] would be like, oh, we'll set up a patient for CPAP. And then they would ask me to write a prescription and I'll do that and fax it back to them." [PCP 6]<br>"Are they all equally useful if you get an appliance from this person or that person? That would be something that I would like to learn more about." [PCP 8] | Diagnosing OSA, Initial Treatment for OSA, Ongoing OSA Management |

Another theme centered on professional relationships and confidence. This theme was most evident during the diagnosis phase, where PCPs positioned themselves as key gatekeepers, drawing on their skills to initiate screening and coordinate referrals. However, confidence and role clarity weakened when initiating treatment and providing ongoing management. While many expressed a strong commitment to providing holistic and longitudinal care, including ongoing symptom management and lifestyle counselling, they often considered certain treatment modalities, such as oral appliances or CPAP titration, to be beyond the scope of primary care. This ambiguity regarding responsibility sometimes resulted in fragmentation of care, where providers were unsure whether follow-up and management should remain with the PCP or be assumed by other care providers.

Provider roles and responsibilities partially offset these challenges. Many PCPs emphasized the therapeutic value of their patient relationships, noting that trust and rapport facilitated discussions about diagnosis and supported treatment adherence. In turn, this reinforced PCPs’ confidence in the impact of their care. PCPs also reflected on their relationships with sleep specialists and respiratory homecare companies, describing collaborative arrangements that enabled them to fulfill their roles despite knowledge gaps or resource limitations. These mutualistic relationships, however, also revealed opportunities for clearer care pathways, stronger interprofessional collaboration, and structural supports to enable more robust primary care engagement in OSA.

## DISCUSSION

In this study, we used a Journey Map to characterize the key steps followed by PCPs in diagnosing and managing OSA. We found that PCPs in Calgary view the Learning, Patient Encounter, and Diagnosing OSA phases favourably, reflected by feelings of confidence, trust, and empowerment. However, frustration, overwhelm, and uncertainty emerge during the Initial Treatment and Ongoing Management stages, leading to negative associated emotions. To our knowledge, this is the first qualitative study to comprehensively map the OSA care journey in primary care from the provider perspective and provides a foundation to identify opportunities for meaningful and sustainable support for PCP-led OSA management.

Our analysis also identified several factors influencing PCP engagement in OSA management. Time pressures and knowledge gaps related to diagnosis and treatment, funding mechanisms, and system navigation were common challenges early in the care process, whereas uncertainties regarding professional roles and pessimism about long-term patient adherence became more prominent in later phases. PCPs consistently valued practical clinical tools and care pathways, timely availability of diagnostic testing, and collaboration with sleep specialists. However, they expressed varying levels of confidence in titrating CPAP therapy and were generally less confident in recommending other OSA treatment modalities such as dental appliances. These findings highlight the need to develop and implement specific care models and supports for role clarity and interprofessional integration at different points in the care journey. However, such strategies must be carefully designed to avoid exacerbating the existing pressures faced by PCPs in already overburdened primary care environments [27, 45].

This study builds on previous literature examining PCP behaviours and experiences with OSA care. Many of the subthemes we identified, such as limited OSA teaching in medical education, inequitable access to funding for OSA treatments, and the perception that the primary role of PCPs is as diagnosticians within the OSA care journey align with prior findings [27, 46, 47]. Barriers such as time pressures, cognitive workload, and questions of professional role identity have also been reported in the primary care management of other chronic conditions, including diabetes and hypertension [48, 49]. Importantly, PCP engagement in OSA care appeared to be shaped not only by knowledge and resource constraints, but also by cumulative emotional experiences from clinical practice. Thus, many of the challenges we identified may reflect broader systemic and behavioural issues within primary care, rather than factors that are specific to OSA management. By mapping TDF domains to the OSA care journey, our study offers insight into how these behavioural and emotional drivers shape PCP engagement.

Several strategies have been proposed to address access and capacity challenges in OSA management, including alternative care provider models, hub-and-spoke systems, and telemonitoring of PAP therapy [21, 50, 51]. By identifying key behavioural factors at specific OSA care journey phases, our findings help determine which of these strategies may be most effective at different steps in the care continuum. Tools such as service blueprints can be used to guide such improvement initiatives by visually mapping actions, roles, and interactions within each phase [52]. Moreover, such tools can enable structured approaches to scaling successful interventions. Importantly, co-design and sustainable implementation of strategies to improve access to high quality OSA care requires strong engagement from all groups involved in OSA care, including patients.

This study has several strengths, including its application of theory-driven behavioural analysis and its comprehensive mapping of the full OSA care journey from the perspective of PCPs, including associated emotions at each phase. The sample included PCPs with varying years of experience from a large urban centre that is demographically diverse. However, there are important limitations that must be acknowledged. First, interviews were primarily designed to understand the impact of a clinical management pathway; thus, while we aimed to include an exploration of perspectives on primary care OSA management more generally, we may not have comprehensively captured barriers and enablers at each phase of the patient care journey. Second, as this study was conducted in Calgary, its findings may have limited transferability to other practice contexts. Nevertheless, the results are consistent with findings from prior studies [27, 46, 47] and we believe are still relevant to other jurisdictions where limited access to specialists necessitates innovative approaches to support timely OSA care [31, 53–55]. Third, the absence of patient perspectives makes it difficult to determine how changes in PCP behaviour would influence patient behaviour and outcomes. Future studies should focus on creating a patient-centered Journey Map to identify overlapping targets for behaviour change.

## CONCLUSIONS

This study identified key behavioural factors that shape PCP decision-making across pivotal stages of OSA care delivery. The Journey Map highlights that while PCPs feel confident and satisfied with diagnosing OSA, they experience increasing challenges and negative emotions as care progresses to treatment and long-term management. These insights can inform potential implementation strategies to better support PCPs in delivering high-quality, primary care-led OSA management.

## Data Availability

The data underlying this article cannot be shared publicly for the privacy of individuals that participated in the study. The data will be shared on reasonable request to the corresponding author.

## Appendix 1 OSA Pathway Project Interview Guide

OSA Pathway Project ◊ **Interview Guide**

### Primary Care Providers

*Use this interview guide with:*

- Primary care providers (PCPs)
  - Family physicians
  - Nurse practitioners
- PCPs who serve Calgary-area patients with OSA
- PCPs who refer OSA patients to specialist or private care providers
- PCPs who use the sleep apnea enhanced primary care pathway, or are aware of it

*Navigating this guide:*

- Overarching domains are set off as section heads and set in all caps
- Key questions in each domain are marked numerically (1., 2., etc.) and in **bold**
- Optional sub-questions and probes for each key question are marked alphabetically (a., b., etc.)
- Deeper prompts, follow-ups, and examples are marked in lowercase roman (i., ii., etc.)

Note that **only key questions** (numbered and in bold) are required to be asked.

Probes and prompts are optional and may be used by the interviewer at their discretion to:

- Elicit more detailed responses
- Track interview progress
- Respond to questions for clarification or requests for examples

Obtain informed consent before proceeding with the interview:

- Interviews will be audio recorded and transcribed verbatim
- Ask permission before starting the audio recording
- Clearly state when audio recording has started and stopped

#### PARTICIPANT BACKGROUND

1. **What is your role?**
  a. Family physician, nurse practitioner, other
  b. How many years have you been in practice?
  c. Do you have any sleep-specific training? (e.g., sleep specialization, sleep-related CME, etc.)
2. **Where do you currently practice?**
  a. *Location:* urban, rural, remote
  b. *Type:* public or private, type of clinic, part of PCN
3. **Can you tell us a bit about the nature of your practice and patient population?**
  a. Approximately how many patients do you have?
    i. On average, about how many patients do you see per week?
    ii. About how many of your patients have an OSA diagnosis?
    iii. About how many of your patients have OSA suspected or probable?
  b. How would you characterize your overall patient population?
    i. *In terms of:* demographics, SES, social determinants of health, etc.
  c. How would you characterize your OSA patients?
    i. *In terms of:* severity, complexity, comorbidities, etc.

#### PATHWAY USE & EXPERIENCE

The Sleep Apnea Enhanced Primary Care Pathway was launched in the Calgary Zone on December 1, 2018.

Have you heard of the pathway? Have you used it?

**4. What is your overall opinion of the pathway?**
  a. *In terms of:* content, quality, usefulness, format, etc.
**5. How did you first hear about the pathway?**
**6. On average, how often do you use the pathway?**
  a. What parts of the pathway do you use most? Why?
  b. What parts do you use least? Why?
**7. What parts of the pathway work well for you? Why?**
  a. *e.g.,* resources, screening tools, navigation tools, Specialist Link, etc.
**8. What parts of the pathway do not work well for you? Why?**
  a. *e.g.,* resources, screening tools, navigation tools, Specialist Link, etc.
**9. Has the pathway impacted your practice or the way you work?**
  a. *If yes,* can you describe the impact?
  b. *If no,* why do you think this is?
**10. We’re curious if the pathway has impacted you in any of these areas:**
  a. Your level of knowledge and confidence <u>managing</u> OSA
  b. Your ability and confidence <u>navigating</u> the system of OSA care
  c. Your ability and confidence <u>communicating</u> with OSA <u>patients</u>
  d. Your ability and confidence <u>communicating</u> with other <u>providers</u> about OSA (e.g., sleep or respiratory specialists, private respiratory homecare companies, other PCPs, etc.)
  e. Your overall workload and patient care?

#### COMMUNICATION

We would like to know a bit more about how you interact and communicate with:

- Sleep specialists
- Respiratory homecare companies (private CPAP providers)

**11. How often do you interact or communicate with:**
  a. Sleep specialists?
  b. Respiratory homecare companies (private CPAP providers)?
**12. What do you primarily interact or communicate with them about?**
  a. *Sample Probes:*
    i. Refer for testing
    ii. Refer for treatment
    iii. Consult without referral
    iv. Follow-up on behalf of patient
**13. Have you used Specialist Link for sleep questions?**
  a. *If yes:*
    i. About how often?
    ii. For what types of questions?
    iii. How was your experience?
  b. *If no:*
    i. Why not? (e.g., not needed, not accessible enough, not clear how to use, etc.)
**14. How would you describe your role in these interactions and communications?**
  a. *Sample Probes:*
    i. Do you have a sense of what your role should be?
    ii. Do you feel your role is clear?
    iii. Does the pathway help you understand your role in these interactions/communications?

#### PATHWAY & SYSTEM IMPROVEMENT

**15. Has the pathway helped overcome any barriers to optimal OSA care?** (for you, or in general)
  a. *If yes:* which barriers, and how?
  b. *If no:* which barriers, and how has it failed?
**16. We identified some common barriers during a previous stakeholder consultation. Do you think the pathway has helped with any of these?**
  a. Wait times, access to care (esp for rural)
  b. System navigation
  c. Role clarity
  d. Conflict of interest
  e. Integration of providers
  f. Use of technology
**17. How can OSA care be further improved?**
**18. How can the pathway be further improved?**
  a. *Sample Prompts:*
    i. Mobile app
    ii. Decision support tools
    iii. More/different educational resources
    iv. Sample case studies
    v. Accompanying CME materials
    vi. Associated CME events/courses

Thank you for taking the time to help us with this work. Before we conclude: is there anything you would like to add – or any suggestions or recommendations you would like to make – that we have not asked you about?

## Appendix 2 COM-B and Theoretical Domains Framework

| **Domains** | **Definition** | **Theoretical constructs represented within each domain** |
| --- | --- | --- |
| Knowledge | An awareness of the existence of something. | Knowledge (including knowledge of condition/scientific rationale, procedures, and task environment)<br>Schemas, mindsets, and illness representations |
| Skills | An ability or proficiency acquired through practice. | Skills (including interpersonal skills and coping strategies)<br>Practice and skills development<br>Competence, ability, skill assessment |
| Memory, attention and decision processes | The ability to retain information, focus selectively on aspects of the environment and choose between 2 or more alternatives. | Memory<br>Attention and Attention control<br>Decision making |
| Behavioral regulation | Anything aimed at managing or changing objectively observed or measured actions. | Self-monitoring and feedback<br>Breaking habit<br>Action planning |
| Beliefs about capabilities | Acceptance of the truth, reality, or validity about an ability, talent, or facility that a person can put to constructive use. | Self-confidence and professional confidence<br>Self-efficacy and self-esteem<br>Perceived behavioral control and competence<br>Beliefs<br>Empowerment |
| Social/professional role and identity | A coherent set of behaviors and displayed personal qualities of an individual in a social or work setting. | Professional, social, and group identity<br>Professional role, boundaries, and confidence<br>Leadership<br>Organizational commitment |
| Beliefs about consequences | Acceptance of the truth, reality, or validity about outcomes of a behavior in a given situation. | Outcome expectancies and its characteristics<br>Beliefs<br>Anticipated regret and consequents |
| Emotion | A complex reaction pattern, involving experiential, behavioral, and physiological elements, by which the individual attempts to deal with a personally significant matter or event. | Anxiety, depression, stress, fear, burn-out<br>Cognitive overload/tiredness<br>Positive/negative affect |
| Goals (I want to) | Mental representation of outcomes or end states that an individual want to achieve. | Goals (distal/proximal, autonomous/controlled)<br>Goal priority<br>Goal/target setting<br>Action planning<br>Implementation intention |
| Intentions (I plan to) | A conscious decision to perform a | Stability of intentions |
|  | behavior or a resolve to act in a certain way. | • Stages of change model<br>• Transtheoretical model and stages of change |
| Reinforcement | Increasing the probability of a response by arranging a dependent relationship or contingency, between the response and a given stimulus. | • Reward (proximal/distal, valued/not valued, probable/improbable)<br>• Reinforcement and incentives<br>• Punishment, sanctions, and consequents<br>• Contingencies |
| Optimism | The confidence that things will happen for the best or that desired goals will be attained. | • Optimism and pessimism<br>• Unrealistic optimism<br>• Identity |
| Environmental context and resources | Any circumstance of a person's situation or environment that discourages or encourages the development of skills and abilities, independence, social competence, and adaptive behavior. | • Environmental stressors<br>• Resources including material resources<br>• Barriers and facilitators<br>• Organizational culture and climate<br>• Person and environment interaction<br>• Salient event and critical incidents |
| Social influences | Those interpersonal processes that can cause individuals to change their thoughts, feelings, or behaviors. | • Social pressure, norms, support, and comparison<br>• Group conformity identity, and norms<br>• Intergroup conflict<br>• Power<br>• Alienation<br>• Modeling |

